# Assessment of the pathogenicity of *Candidatus* Rickettsia colombiensis in a Syrian hamster (*Mesocricetus auratus*) model and serological cross-reactivity between spotted fever species

**DOI:** 10.1101/2025.05.06.25327046

**Authors:** Jorge Miranda, Alejandra García, Cristina Cervera-Acevedo, Sonia Santibañez, Aránzazu Portillo, José Oteo, Salim Mattar

**Affiliations:** Instituto de Investigaciones Biológicas del Trópico, Universidad de Córdoba, Córdoba, Colombia; Centre of Rickettsiosis and Arthropod-Borne Diseases (CRETAV), Department of Infectious Diseases, San Pedro University Hospital – Centre for Biomedical Research (CIBIR), La Rioja, Logroño, Spain

**Keywords:** Pathogenic, cross-reactivity, *Rickettsia*, Indirect Fluorescent Antibody Technique

## Abstract

**Background:** *Rickettsia* are obligate intracellular gram-negative bacteria of the class Alphaproteobacteria and are vector-borne species that cause diseases in humans and animals. New *Rickettsia* species have been involved as human pathogens; however, the pathogenic potential of other species is unknown. *Candidatus* Rickettsia colombiensis is a new species in the spotted fever group of unknown pathogenicity that is phylogenetically related to other pathogenic species.

**Objetive:** To evaluate the pathogenic potential of *Ca*. R. colombiensis in Syrian hamsters (*Mesocricetus auratus*) and analyzed its cross-reactivity against other SFG rickettsia species in human and Syrian hamster sera.

**Methods:** *Ca*. R. colombiensis was isolated from *Amblyomma dissimile* using a shell vial. Subsequently, five male hamsters were inoculated intraperitoneally (IP) and five intradermally (ID) with 1 × 106 Vero cells infected with *Ca*. R. colombiensis. One control animal was used in each group of the study. The health status of the animals was assessed daily, and necropsies were performed on days 5, 10, 15, and 16 DPI. Serum samples for indirect immunofluorescence and tissues were processed for qPCR and immunohistochemistry.

**Results:** All animals remained healthy during the trial and showed no changes in their physiological parameters. No Rickettsia DNA was detected in any of the tissues. Animals infected with *Ca*. R. colombiensis showed IgG antibody titers ranging from 1:64 to 1:1024. The control animals were negative. Regarding human sera, 56% (84/150) had IgG antibodies against Ca. R. colombiensis antigen. Sera with titers equal to or higher than 1:64 were 100% seroreactive.

**Conclusions:** *Ca*. R. colombiensis caused a subclinical infection in hamsters, suggesting the possibility of infecting other mammals. However, the clinical, pathological, and molecular findings are inconclusive in establishing that *Ca.* R. colombiensis is a pathogenic species. Regarding cross-reactivity, it is possible to serologically diagnose Rickettsial infection using *Ca*. R. colombiensis as antigens.

**Author summary:** Rickettsiosis is a human and animal disease caused by the bacterial Rickettsia species, which are mainly transmitted by arthropods. The clinical spectrum of rickettsiosis ranges from asymptomatic infection to severe and fatal diseases, such as Rocky Mountain spotted fever caused by Rickettsia rickettsii. In recent years, the number of species belonging to the genus Rickettsia has increased, in part due to the progress in the detection and identification of these agents by the application of modern molecular techniques. Much of this new *Rickettsia* species, known as Candidatus, has been involved as animal and human pathogens; however, for other Candidatus Rickettsia species, the ability to cause diseases is unknown. *Candidatus* Rickettsia colombiensis is a new species phylogenetically related to pathogenic species. Here, we evaluated, for the first time, the pathogenic potential of this microorganism in a hamster animal model and analyzed its cross-reactivity against other members of the Rickettsia genus in human and hamster sera to determine the possibility of using *Candidatus* Rickettsia colombiensis as antigens to diagnose rickettsiosis in regions where immunologic reagents are scarce and high biosafety level laboratories do not exist. In this report, we demonstrate that *Candidatus* Rickettsia colombiensis causes an inapparent infection in hamsters and cross-reactivity between other members of the Rickettsia genus.

## Introduction

*Rickettsia* are obligate intracellular gram-negative bacteria of the class Alphaproteobacteria and are vector-borne species that cause diseases in humans and animals. They are globally distributed and are a considerable public health concern [1]. Pathogenic rickettsiae are associated with hematophagous arthropods, such as ticks, mites, fleas, and lice, which act as transmission vectors and reservoirs in their natural life cycle. However, Rickettsiae spp. have also been found in various hosts, including herbivorous arthropods, leeches, and amoebae [2,3].

Based on phylogeny, clinical symptoms, and antigenic properties, rickettsiae are classified into three pathogenic groups: spotted fever (SFG), typhus (TG), and transitional (TRG) groups. In addition, a group of non-pathogenic rickettsiae is poorly studied compared to the pathogenic rickettsiae groups called the ancestral group (AG) [4,5]. TG comprises two species, *Rickettsia prowazekii* (louse-borne epidemic typhus) and *Rickettsia typhi* (murine typhus), which are transmitted by lice and infected flea feces, respectively [4,6]. TRG includes *Rickettsia felis* (flea-borne spotted fever), *Rickettsia australis* (tick-borne Queensland tick typhus), and *Rickettsia akari* (mite-borne rickettsialpox), which are mainly transmitted by fleas [7]. *R. bellii* and *Rickettsia canadensis* are considered non-pathogenic and endosymbionts of ticks and belong to AG [8]. The SFG contains the most significant number of rickettsiae, with more than 20 recognized species, including lethal ones such as *Rickettsia rickettsii* (Rocky Mountain spotted fever) and *Rickettsia conorii* (Mediterranean spotted fever) [4,6,9].

Many Rickettsia species discovered in ticks were initially described as endosymbionts because they were not associated with diseases in animals or humans [10]. However, some pathogens, such as *Rickettsia parkeri* and *Rickettsia slovaca*, were later identified as pathogens in humans and animals based on clinical records and microbiological approaches indicating pathological findings [11,12].

New *Rickettsia* species have been involved as human pathogens as occurred several years ago with *Candidatus* Rickettsia rioja [13] or recently with *Candidatus* Rickettsia lanei, reported in two patients with Rocky Mountain spotted fever (RMSF-like) in California, USA [14]. This emergence of new pathogens, coupled with climate change that causes the expansion of vectors and the low interest in the study of rickettsiosis, which is considered a neglected disease, could put half of the world’s population at risk of contracting rickettsiosis from a species of SFG [15,16].

In 2012, a new species of Rickettsia, *Candidatus* Rickettsia colombiensis (formerly *Candidatus* Rickettsia colombianensis), was described for the first time. Rickettsia was detected in *Amblyomma dissimile* ticks collected from *Iguana iguana* in Córdoba, Colombia [17]. *Ca*. R. colombiensis has the *ompA* gene, which is located in the SFG and is phylogenetically related to *Rickettsia tamurae* and *Rickettsia monacensis*, both of which are recognized as human pathogens [18,19].

In 2020, the names of bacteria in the Candidatus status were published. The list was created to correct Candidatus names and comply with the current requirements of the International Code of Nomenclature of Prokaryotes. Considering these requirements, *Candidatus* Rickettsia colombianensis was renamed *Candidatus* Rickettsia colombiensis [20]. *Candidatus* Rickettsia colombiensis has been reported in ticks of the genera *Amblyomma*, *Rhipicephalus*, and *Dermacentor* sp. in different parts of Colombia [21–25]. It has also been detected in other countries in Central and South America [26–30]. In Colombia, the main vector of *Ca*. R. colombiensis is *A. dissimile*, with infection rates between 15% and 100% [17,25,31,32]. Given the high infection rate of this rickettsia, there is a risk of transmission to humans and animals when bitten by this tick species. All Rickettsia species are thought to have the potential to be pathogenic in vertebrates. The limiting factor for disease is the opportunity for transmission to vertebrate hosts [10,33], however, the pathogenicity of *Ca.* R. colombiensis in humans and animals is unknown.

Serological cross-reactivity between rickettsia species of the same and different groups has been well documented [34]. *Ca*. R. colombiensis is a member of the SFG, so it would have cross-reactivity with other SFG members that circulate in the region and cause infections in humans and animals. This cross-reactivity could provide an opportunity for diagnosis in areas where laboratory supplies are scarce. In this regard, slides with *Ca.* R. colombiensis may aid in the diagnosis of rickettsiosis caused by SFG rickettsiae.

This study aimed to evaluate the pathogenic potential of *Ca*. R. colombiensis in Syrian hamsters (*M. auratus*) and analyze cross-reactivity against other rickettsial species of the spotted fever group in sera from humans and Syrian hamsters.

## Methods

### Study type and tick collection

In 2023, a prospective descriptive study was conducted in the Department of Córdoba. *Amblyomma dissimile* ticks were collected from iguanas (*Iguana iguana*) and snakes (*Boa constrictor*). The ticks were kept alive and transported to the Institute of Biological Research of the Tropics Laboratory, where they were stored at –90°C.

### Isolation of *Candidatus* Rickettsia colombiensis

Ten *A. dissimile* specimens were cultured using the shell vial technique to isolate *Ca*. R. colombiensis. The ticks were disinfected with iodine alcohol for 10 min, washed with sterile water, and macerated in Brain Heart Infusion (BHI) broth (Thermo Scientific™ Oxoid™). Macerate (200 µL) was inoculated into shell vials containing Vero cells and supplemented with Dulbecco’s Modified Eagle’s medium (DMEM) containing 5% bovine fetal serum (BFS) enriched with iron, 1% penicillin-streptomycin, and amphotericin B and incubated at 28°C and 32°C. The presence of *Rickettsia* was determined using *Giménez* staining and PCR [35,36]. The positive shell vials were transferred to a 12 cm^2^ bottle, and once 90% infection was reached, the cells were transferred to a 75 cm^2^ flask. When >90% infection was obtained, the culture was used to infect Syrian hamsters and develop immunofluorescence assay (IFA) slides. One part was cryopreserved in sucrose-phosphate-glutamate (SPG) buffer at –90°C [36,37].

### Evaluation of *Candidatus* Rickettsia colombiensis pathogenesis in an experimental model of Syrian hamster (*Mesocricetus auratus*)

Twelve male hamsters aged 6 weeks and specific pathogen-free (SPF) were used. Ten animals were included in the experimental group and two in the control group. The method of infection and evaluation of hamsters was modified according to previously described methods [38]. Anesthetized animals were inoculated intraperitoneally (day 0) in five hamsters intraperitoneally (IP group) with 500 µL of 1 × 10^6^ Vero cells infected with *Ca*. R. colombiensis strain AdCor 5 third passage and five hamsters intradermally (ID group) with 1 ml of the same concentration of infected Vero cells with *Ca*. R. colombiensis in DMEM medium. Control hamsters (one for IP and one for ID) were inoculated similarly to uninfected Vero cells. A flowchart of the infection and euthanasia process is shown in Figure 1. The hamsters were analyzed daily for physiological parameters, including weight (g), temperature (°C), and mortality. Qualitative parameters were also analyzed, including the appearance of feces, urine, hair, skin, hydration level, behavioral changes, oral mucosa, and the conjunctiva. The appearance of scrotal reactions, such as edema, congestion, and necrosis, typical of acute *Rickettsia* infection in these animals, was also evaluated [39,40]. On days 5, 10, 15, and 16 DPI, the hamsters were sacrificed, and blood and serum samples were collected for indirect immunofluorescence (IFA) tests. The sera were analyzed using various Rickettsia antigens, including *Ca.* R. colombiensis, *R. rickettsii* and *R. parkeri*. The organs were preserved, and qPCR and immunohistochemistry were performed on them. The euthanasia procedure is illustrated in Fig 1. The controls in each group were euthanized at 16 DPI. The following organs were collected at necropsy: brain, heart, lungs, spleen, liver, kidneys, and blood vessel fragments. Some of these tissues were frozen at −90°C for PCR analysis and culture. The remaining tissues were used for histopathological studies (10% buffered formalin). Organs were processed using standard histopathological protocols, and sections were prepared using hematoxylin and eosin (H&E).

**Fig 1.**
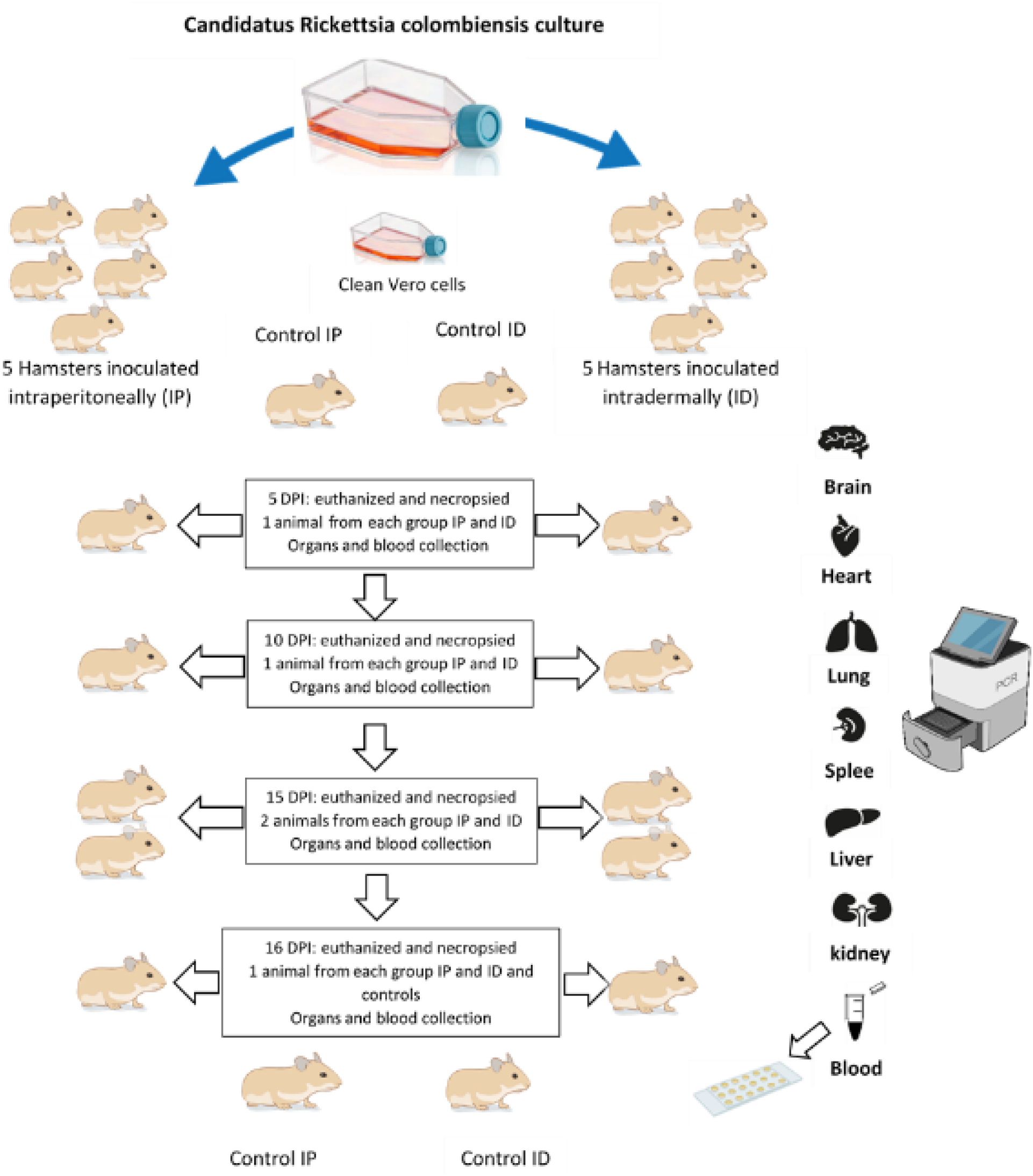
Flowchart of infection and euthanasia of the 12 Syrian hamsters.

### Ethical aspects

The hamsters were anesthetized with an intramuscular mixture of Ketamine and Xylazine (10:1), and a safe dose of the anesthetic combination of 0.1-0.3 ml was used for animals weighing 100-200 grams. Euthanasia was performed using an overdose of sodium pentobarbital, which was calculated according to the weight of the animal. The study was approved by the Institute of Biological Research of the Tropics Ethics Committee of the University of Córdoba Minutes 010-2022 on October 12, 2022. This study was approved by the University of Antioquia Ethics Committee for Animal Experimentation (minutes of session 151, April 11, 2023).

### Preparation of slides and immunofluorescence assays (IFA)

Cells with a level of infection ≥ 90% with *Ca.* R. colombiensis from the 75 cm^2^ bottle was scraped off, transferred to a tube, and centrifuged (13000 g/10 min). The pellet was resuspended in PBS containing 2% bovine fetal serum (BFS) and 0.1% sodium azide. Then, 10ul of the infected cells was placed in each well of the slide and allowed to dry at room temperature in a biosafety cabinet. The antigen was fixed with acetone for 10 min and stored at-80 C until use (Horta, Maurício C. et al., 2004). Serodiagnosis by IFA test remains the gold standard for Rickettsial infections using seroconversion and four-fold antibody titer increases in two paired sera separated by at least 2–6 weeks [41]. Briefly, each well of the slide contained *Ca*. R. colombiensis antigen, was cover with 10 µl of the hamster and human sera diluted 1:64 (10 µl of serum + 630 µl of PBS). The slides were then incubated at 37°C for 30 min. The cells were then washed three times with PBS. Subsequently, 15 µL of conjugate was added; for hamsters, a 1:400 dilution of Goat Anti-Hamster IgG (H+L) and Mouse/Rat ads-FITC was used. Cat. No 6061-02 (SouthernBiotech). For human sera, a 1:400 rabbit anti-human IgG (H+L)-FITC conjugate (Cat. No. MBS524382 (MyBioSource) was incubated at 37°C for 30 min. The slides were washed twice with PBS + Evans Blue (0.3 ml per 100 ml of PBS wash) [42]. The slides were observed under a fluorescence microscope at 400X.

### Cross-reactivity of hamster and human serum

Serological cross-reactions between different Rickettsia species and the same rickettsial group or even with other bacterial genera can occur because of the production of antibodies that recognize proteins with the same antigenicity [43]. IFA test can help in the identification of rickettsial species; if it demonstrates a fourfold higher dilution compared to other rickettsial antigens, this may be suggestive of a causative organism; however, cross-reactions or the maintenance of high antibody titers for a long time can interfere with this technique [44]. To establish the degree of cross-reactivity of the antibodies in the hamster and human sera infected with *Rickettsia* sp. of the spotted fever group, the slides were antigenized with *R. rickettsii*, *R. parkeri*, and *Ca*. R. colombiensis was used. Slides with *R. rickettsii* and *R. parkeri* were donated by Professor Marcelo Labruna of the Universidad São Paulo, Brazil.

### Human sera for cross-reactivity study of Rickettsia sp. in the spotted fever group

A total of 150 serum samples were obtained from individuals diagnosed with *Rickettsia* infection of the spotted fever group from Córdoba, Colombia (Barrera et al., 2015; Hidalgo et al., 2011). Four percent of these subjects (6/150) had confirmed infection with *R. rickettsii* (by PCR and sequencing). The remaining 96% (144/150) of the participants underwent seroprevalence studies with positive IFA results. Slides with *R. rickettsii* antigens were used to detect antibodies against SFG rickettsia in human serum.

### Detection of Rickettsia using qPCR

DNA was extracted from hamster sera, tissues, and cultures using a PureLink™ Genomic DNA Purification Kit (Ref: 1820-00, Invitrogen™). qPCR was performed with a TaqMan probe specific for the Rickettsia genus using primers CS-5 and CS-6 [36]. The assay was performed using FastStart Taq DNA Polymerase (Roche) for molecular characterization of Ca. R. colombiensis isolates. The *OmpA* gene was amplified with primers Rr 190.70 and 190-701 [45]. *Rickettsia amblyommatis* DNA was used as a positive control, and molecular-grade water was used as a negative control. To confirm infection by *Ca*. R. colombiensis, the amplicons were purified with NucleoSpin Gel and PCR Clean Up (Ref: 720609.50, Bioanalysis) and both chains were sequenced using the dideoxy method in a SeqStudio, (Applied Biosystems) at the research Centre of Rickettsiosis and Arthropod-Borne Diseases (CRETAV), Infectious Diseases Department, San Pedro University Hospital – Center for Biomedical Research (La Rioja, Spain).

### Phylogenetic analysis

To confirm the phylogenetic position of *Ca*. R. colombiensis, the *ompA* gene sequence was used, and the analysis was performed using IQ-TREE. The sequences obtained were aligned with other rickettsia sequences available in GenBank. Kimura’s parameter two was used as the best nucleotide substitution model for the phylogenetic trees. The trees were constructed using the maximum likelihood method, and the reliability of the analysis was determined by bootstrap analysis with 1,000 replicates [46].

## Results

### Isolation of *Candidatus* Rickettsia colombiensis

Two strains of *Ca*. R. colombiensis (AdCor5 and AdCor6) were also isolated. The *gltA* gene was detected in both strains using qPCR (primers CS5 and CS-6), and the *ompA* gene (632 bp fragment) was detected using conventional PCR. Assays and phylogenetic analyses were performed using *Ca*. R. colombiensi isolate AdCor5. Sequencing of the amplified *ompA* products showed 100% of nucleotide identity with the R. colombianensis strain (JF905458). Phylogenetic analysis of the partial *ompA* sequence revealed that Ca. R. colombiensis strain AdCor5 was grouped with other reported *Ca.* R. colombiensis sequences detected in Colombia and Brazil at a bootstrap level of 100%. Furthermore, phylogenetic analysis showed that strain AdCor5 was grouped in the same clade as *R. tamurae* and *R. monacensis,* with high bootstrap levels of 99% (Fig 2). The nucleotide sequence of the *ompA* gene fragment of strain AdCor5 was deposited in GenBank under accession number PQ658084.

**Fig 2.**
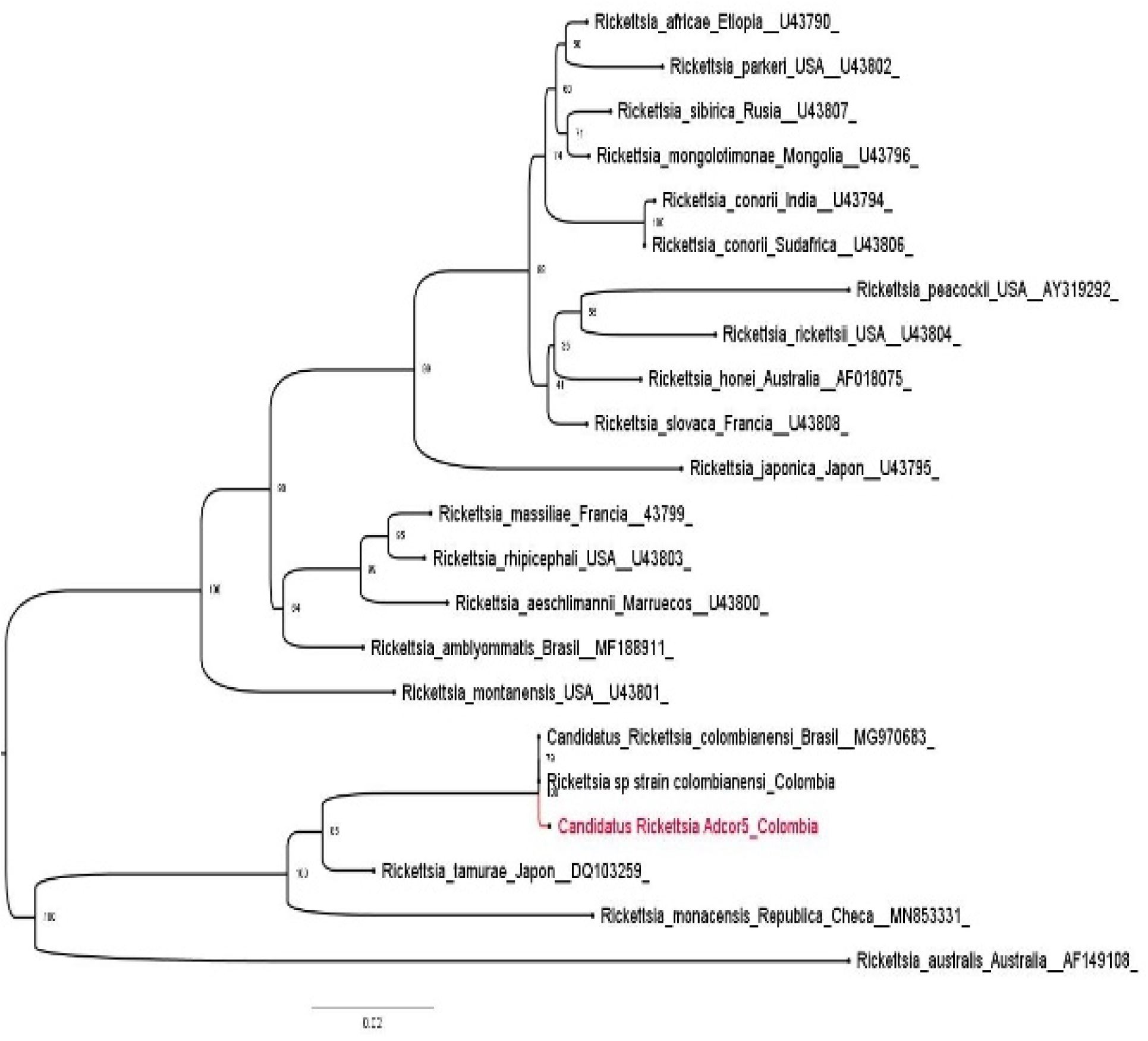
Phylogenetic position of *Ca*. R. colombiensis AdCor5 (in red) among the validated SFG rickettsial species. The maximum likelihood method was based on the Tamura-Nei model, and partially broken sequences were shown. The analysis included 22 nucleotide sequences from the samples. All positions containing gaps and missing data were removed. The final dataset contained 564 positions. Evolutionary analyses were conducted using IQ-TREE.

### Evaluation of the pathogenesis of *Ca*. R. colombiensis in an experimental Syrian hamster model (*M. auratus*)

All animals remained healthy during the 16 days of the trial. They did not show changes in quantitative physiological parameters, such as weight loss, temperature, and mortality. Animals in the IP group showed an average weight gain of 16 g (5.3 – 21 g). The ID group showed an average weight gain of 18.7 g (7 – 19.7 g). Figs 3A and B show the parameters of body weight and daily recorded temperature of the animals (Figure 3 C and D). No abnormal changes were observed in the feces, urine, hair, skin, hydration level, behavior, appearance of the skin, oral mucosa, or conjunctiva, and no reactions such as edema, congestion, or necrosis were observed in the scrotum. The animals in the control group did not exhibit any symptoms of disease or pathological changes. No hemorrhage, vasculitis, or tissue necrosis was observed during necropsy. Histopathological analysis of the liver, lung, brain, spleen, aorta, and kidney tissues stained with hematoxylin and eosin revealed no lesions associated with rickettsial infection. However, some organs, such as the liver and kidney, presented lesions, such as moderate multifocal lymphocytosis in the interstitial tubule, lymphocytosis, focal membranoproliferative glomerulopathy and mild multifocal periportal plasmacytosis in the liver (S1 Fig).

**Fig 3.**
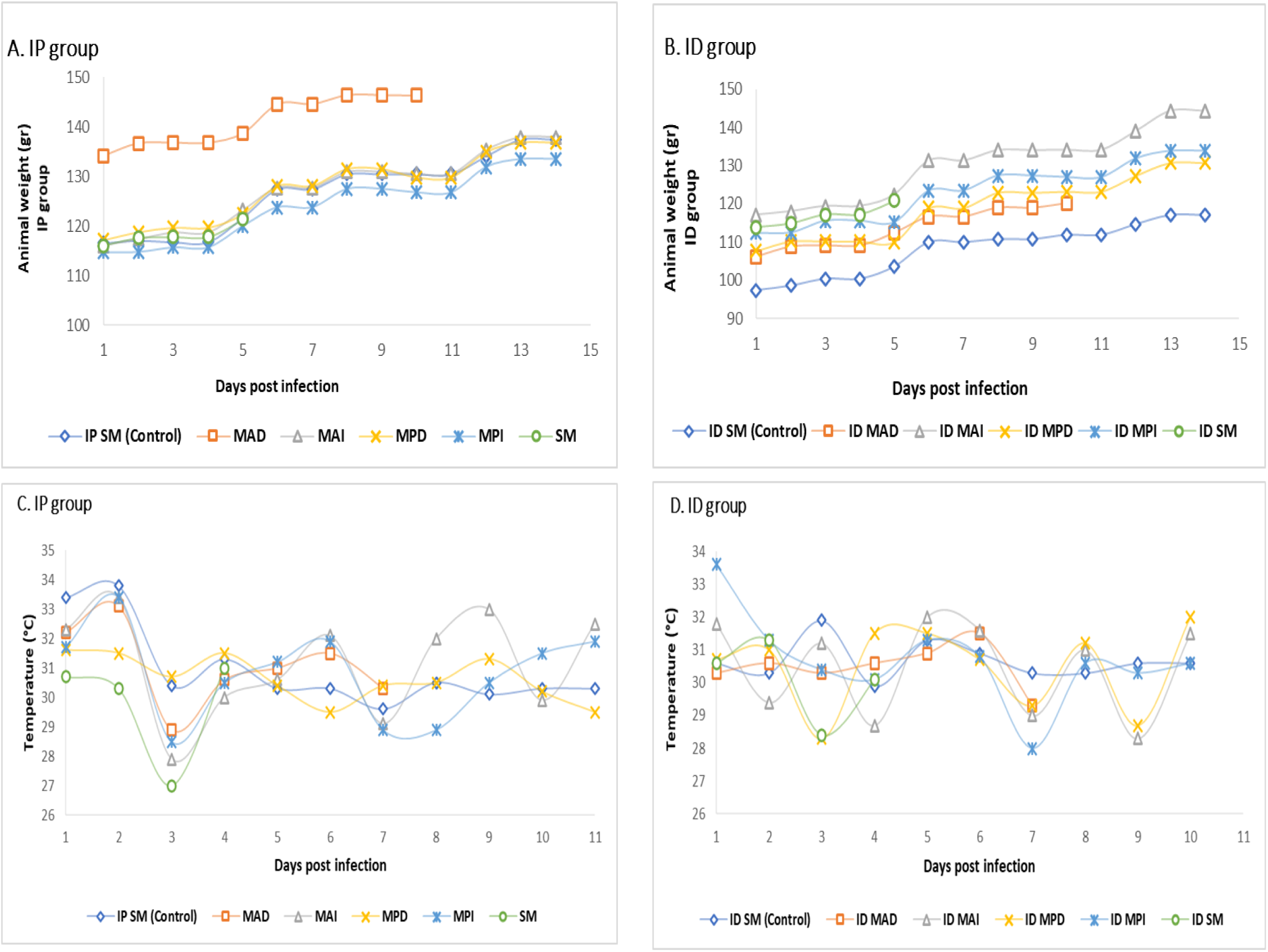
Weight measurements for both groups of *Ca*. R. colombiensis-infected animals. (A and B). Temperature measurements in both groups (C and D). IP: Intraperitoneal, ID: intradermal.

### Detection of Rickettsia in Syrian hamster tissues

Eighty-four DNA extractions were performed from each animal’s blood, serum, liver, lung, brain, spleen, and kidney, all of which were negative for *Rickettsia* sp.

### Indirect immunofluorescence (IFA) was performed on Syrian hamsters

Twelve serum samples from the animals included in this study were analyzed. All sera from animals in the IP and ID groups inoculated with *Ca*. R. colombiensi presented anti-*Ca*. R. colombiensis IgG antibodies were detected from 5 DPI with titers of 1:64. The maximum titer was 1:1024 at 15 days post-inoculation (DPI). The control animals did not contain any antibodies. The antibody titers in each inoculation group are shown in Fig 4.

**Fig 4.**
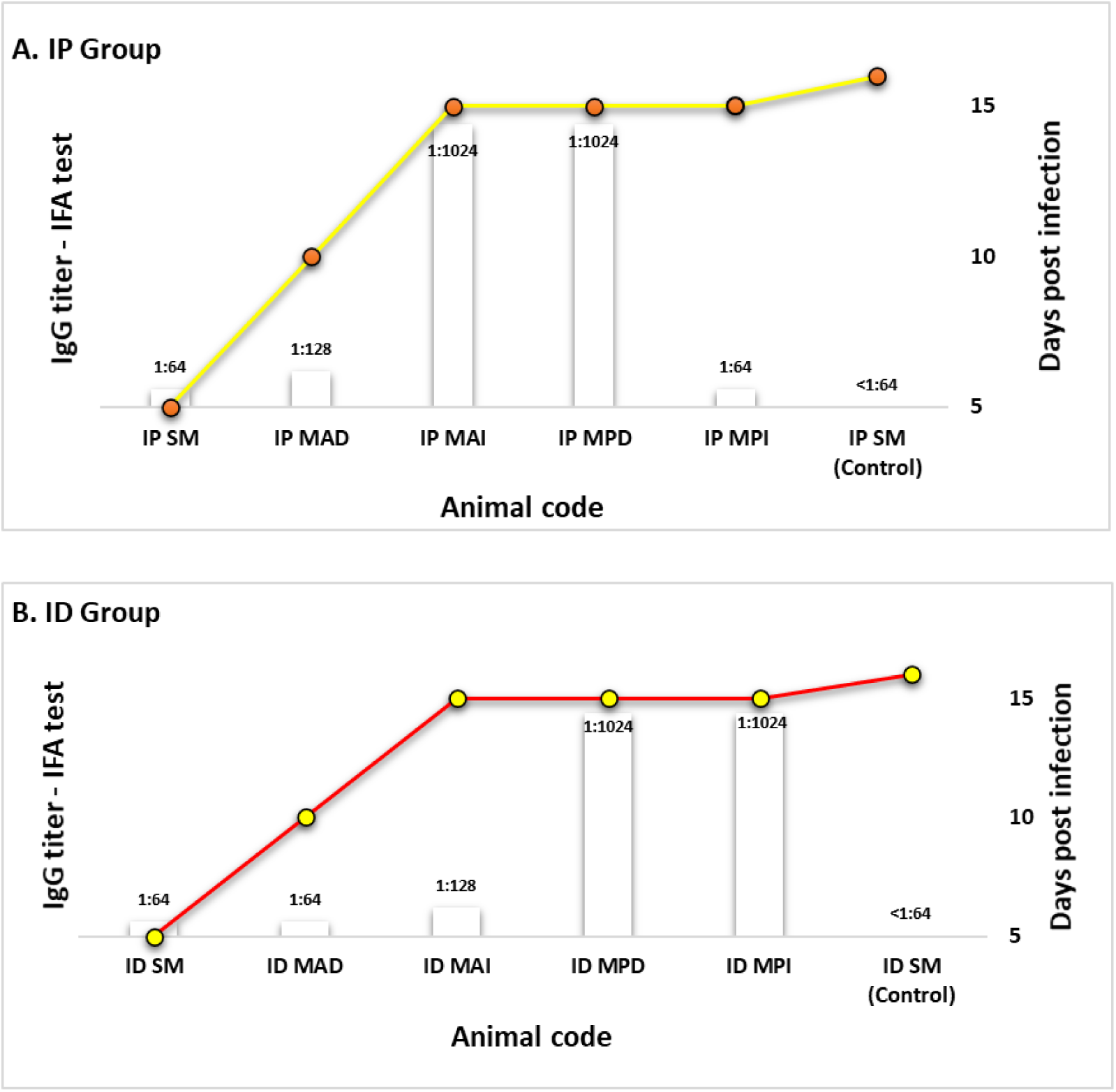
Anti-Rickettsia IgG endpoint titers of Syrian hamster inoculated with *Ca*. R. colombiensis. IP: Intraperitoneal, ID: intradermal.

### Cross-reactivity in hamster and human sera

None of the sera from the 12 *M. auratus* (10 from the IP– and ID-infected animals and two controls from each group) presented antibodies against *R. rickettsii* or *R. parkeri* antigens. All 10 serum samples from the infected animals presented with antibodies against *Ca*. R. colombiensis antigen, with titers between 1:64 – 1:1024. Regarding human sera, 56% (84/150) had IgG antibodies against *Ca*. R. colombiensis antigen, demonstrating cross-reactivity. The same sera were tested for *R. parkeri*, and 54% (81/150) of the sera had antibodies. Fig 5, A shows the number and antibody titers of human sera tested against *R. rickettsii*, *R. parkeri*, and *Ca*. R. colombiensis antigens. However, of the 150 sera tested, 30 were selected with titers equal to or greater than 1:64, distributed as follows: three sera with titers of 1:64, six sera with titers of 1:128, 11 sera with titers of 1:256, nine sera with titers of 1:512, and one serum with a titer of 1:1024. This selection was made because 52% of the sera (79/150) had an antibody titer of 1:64. A low titer of 1:64 did not indicate acute or recent infection; therefore, these patients were excluded. All 30 samples were 100% seroreactive for *Ca*. R. colombiensis antigen. However, the titers of *R. rickettsii* and *Ca*. R. colombiensis were differentially expressed. Figure 5 shows the titers obtained from the analysis of these samples (Fig 5 B).

**Fig 5.**
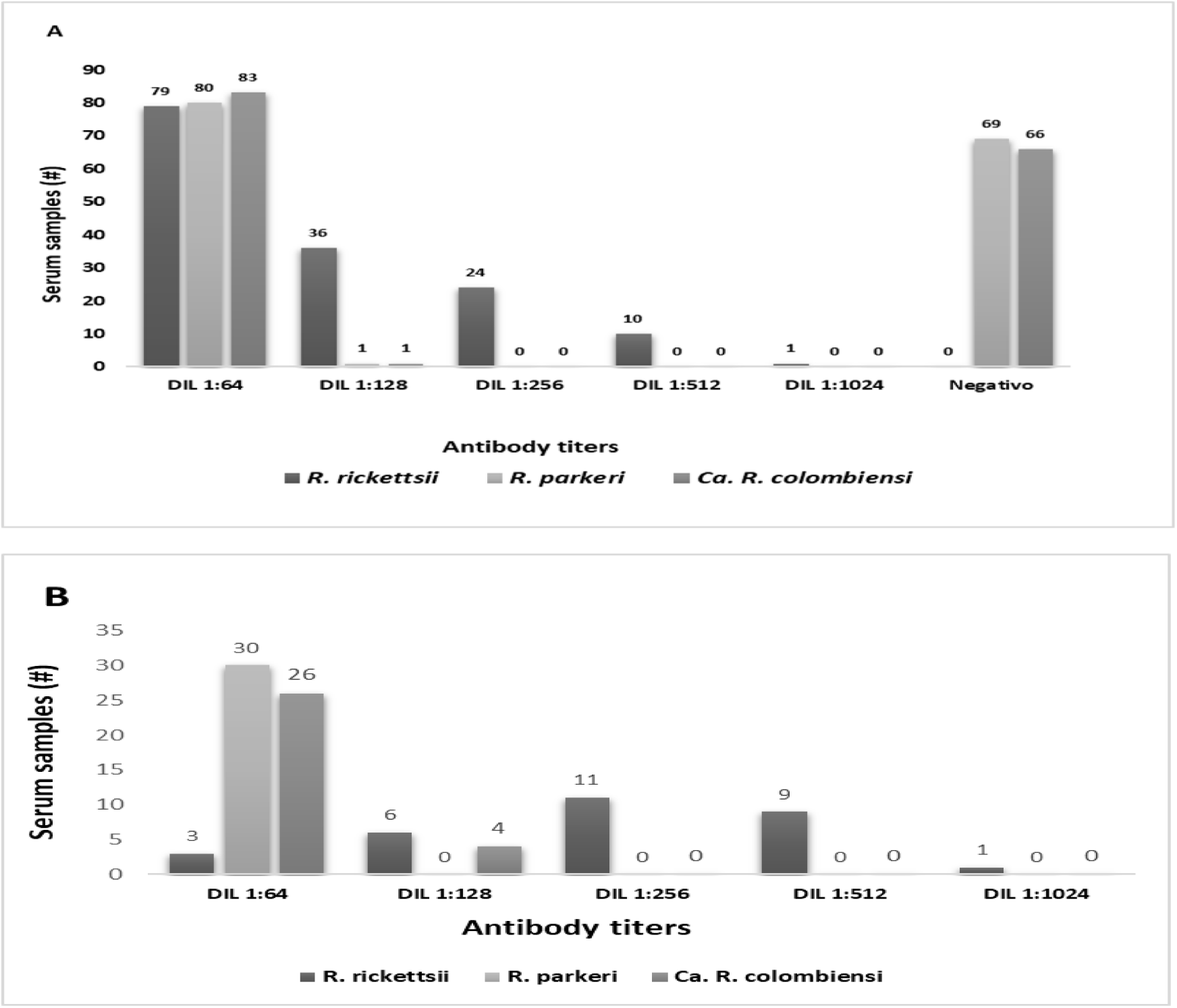
A. Number and antibody titers of human sera tested against R. rickettsii, R. parkeri, and Ca. R. colombiensis antigens in 150 subjects. **B**. Number and antibody titers of human sera from 30 subjects.

## Discussion

In the present study, anti-Ca. R. colombiensis IgG antibodies were detected in Syrian hamsters and cross-reactivity among *Ca*. R. colombiensis and other spotted fever rickettsia group (*R. rickettsii* and *R. parkeri*) was confirmed. The animal model did not show signs of disease caused by pathogenic Rickettsia. However, some animals presented with lesions in the histological sections of the kidney and liver. Since no vasculitis-related rickettsiosis was identified in tissues, these lesions could indicate lesions of a genetic background that are frequent in rodents used for research purposes and are possibly not associated with the pathological process of the infection (S1 Fig) [47].

Animal models can imitate the complex pathogenesis of infection [48]. In this sense, different animal species have been used as models for studying the pathogenesis of Rickettsia; Guinea pigs (*Cavia porcellus*) have been the most widely used [39,49]. C3H/HeN, C57BL/6, Balb/c mice, and Syrian hamsters (*M. auratus*) have been used as models for pathogenesis studies [50–52]. The animal model used in the present study (*M. auratus*) has been reported as one of the best models for pathogenesis studies because of its physiological similarity to the human immune system compared with that of other animal species [48].

Human pathogenic rickettsiae, such as *R. prowazekii* and *R. rickettsii*, cause lethal infections or diseases in guinea pigs [49]. The first studies on pathogenicity proposed the hypothesis that the infection and lethality rates of Rickettsia observed in guinea pigs could be an extrapolated model to assume the possible infection and lethality in humans. However, McDade [53] reported no correlation between animal and human fatalities and suggested that this extrapolation should be performed cautiously.

To our knowledge, this is the first study to evaluate the pathogenicity of *Ca*. R. colombiensis in an animal model. Other Rickettsia species have also been reported to be non-pathogenic in vectors and were subsequently assessed in animal models. In this regard, *Rickettsia amblyommatis* (formerly *Candidatus* Rickettsia amblyommii) was initially classified as non-pathogenic because no signs of disease were found in guinea pigs [54,55]. However, in both studies, an IgG-type immune response was observed, indicating subclinical infection. These results agree with those reported in our study, where no clinical or histopathological signs of the disease were observed, but an evident antibody response was detected. In previous studies on guinea pigs, *R. amblyommatis* was considered non-pathogenic [9,56]. However, recent studies have reported clinical findings such as bilateral enlargement of the testicles and histopathological findings such as inflammatory processes around the testis and necrotic lesions on the liver. In addition, an IgG-type immune response was demonstrated, indicating an infection similar to that caused by pathogenic Rickettsia of the SFG. These contradictory results could be explained by the fact that three different strains of *R. amblyommatis* were used, isolated from other sites in the United States: strain WB-8-2T [55], strain North Texas isolates [54], strain Lake Alexander [9], and strain 9-CC-3-1 isolated from Costa Rica [56].

Another Rickettsia species of unknown or controversial pathogenicity that has been evaluated in animal models to determine its pathogenicity is *Rickettsia bellii*. This species is considered non-pathogenic. The concept of nonpathogenicity is based on the fact that it does not cause disease in guinea pigs or *Microtus pennsylvanicus* [57]. These results are similar to those of another study in which no clinical symptoms were observed in guinea pigs or opossums (*Didelphis aurita*) infected with *R. bellii* [58]. However, high titers of anti-*R. bellii* antibodies. These data are consistent with those reported in the present study for *Ca*. R. colombiensis did not produce clinical signs but did produce an immunological response, indicating subclinical infection.

*Rickettsia monacensis* was initially detected only in *Ixodes ricinus* in several European countries. In an animal model of six Syrian hamsters injected intraperitoneally with *R. monacensis*, all the animals remained healthy during the study. However, five seroconverted isolates with a high titer of 1:16,384 demonstrated subclinical infection [52]. These results are similar to those of the present study, in which we used the same animal model, but none of the animals presented with symptoms. Nevertheless, the animals developed an immune response against *Ca*. R. colombiensis antigen. It is important to note that currently, *R. monacensis* is recognized as a human pathogen [59].

The results of the present study demonstrate that antibodies induced by pathogenic rickettsiae of the SFG (*R. rickettsii* and *R. parkeri*) have 100% cross-reactivity with the antigens of *Ca*. R. colombiensis when the antibody titers were higher than 1:128. The cross-reactions observed in the present study were similar to those found in human sera using IFA in the USA [60]. In that study, it was found that all sera had a cross-reaction among the species *R. rickettsii*, *R. montanensis*, *R. parkeri*, and *R. amblyommatis*. Serum cross-adsorption and western blotting results revealed that 35 serum samples had specific antibodies against *R. amblyommatis*, 1 against *R. montanensis*, and 1 against *R. parkeri*, and none of the samples had specific antibodies against *R. rickettsii*. The cross-reactions in our study were similar to those of another study conducted in Colombia in people living in an endemic area of rickettsiosis [61]. In that study, a cumulative incidence of 6.23% (17/273) was reported after one year of follow-up. IFA was performed with antigens from *R. parkeri*, *R. rickettsii*, *R. amblyommatis*, *R. belli*, *R. rhipicephali*, and *R. felis* to establish the species of rickettsia involved. However, it was impossible to establish the probable species responsible for human infection because all 17 subjects had cross-reactivity against the six antigens used. Only three participants showed titers higher than 1:1024 (two participants with a titer of 1:2048 for *R. rickettsii* and *R. amblyommatis* and one participant with a titer of 1:1024 for *R. rickettsii* and *R. amblyommatis*) (Table 1).

**Table 1.**
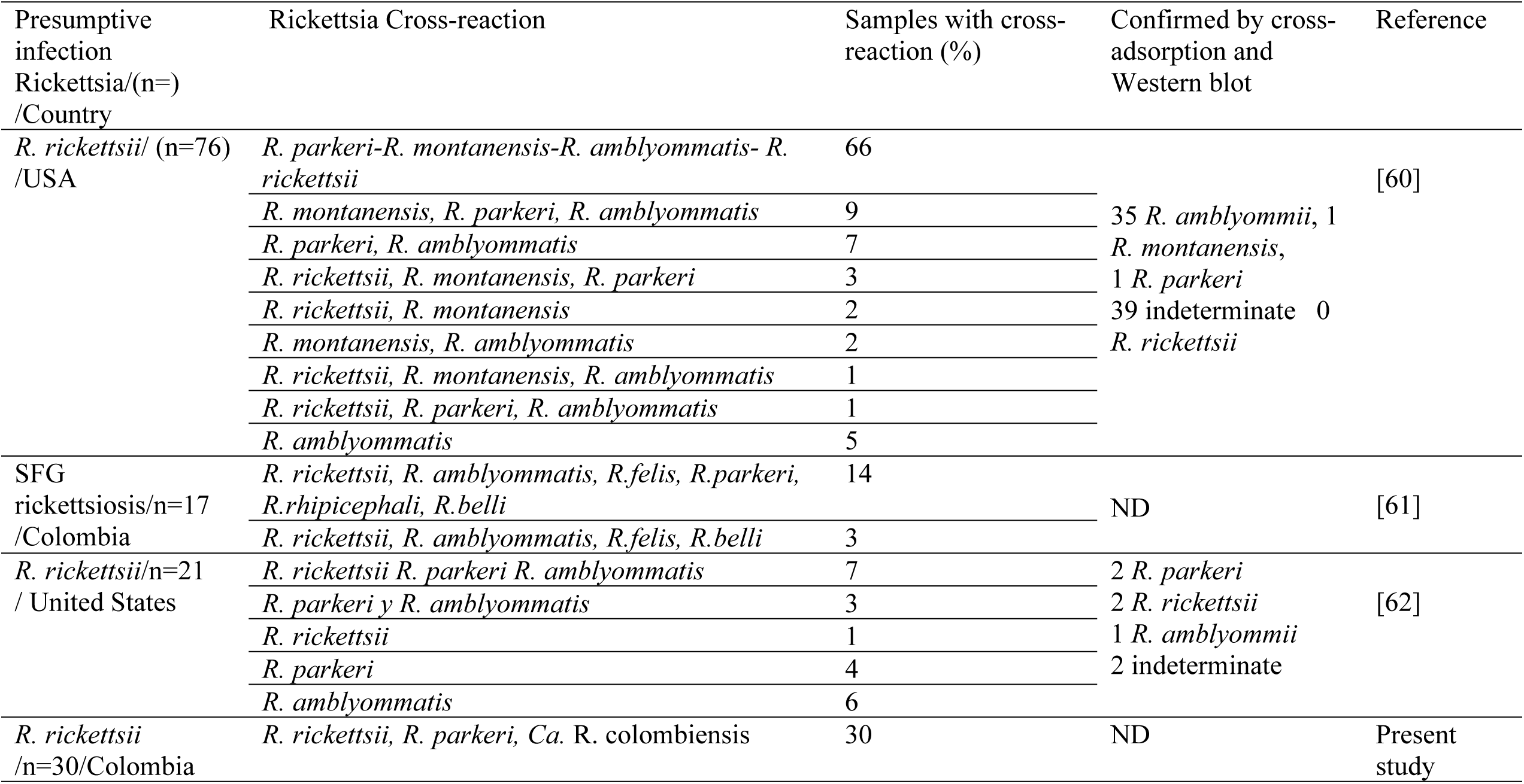
Serological reaction studies between SFG species in different sites in South and North America.

Another study that evaluated cross-reactivity in paired sera presumptively diagnosed with Rocky Mountain spotted fever (RMSF) against *R. parkeri* and *R. amblyommatis* antigens [62], found that 21 patients seroconverted and showed cross-reactivity between the three species tested. Regarding seroconversion, seven participants seroconverted to all three antigens, three participants seroconverted to two antigens (*R. parkeri* and *R. amblyommatis*), and 11 participants seroconverted to a single antigen (one to *R. rickettsii*, four to *R. parkeri*, and six to *R. amblyommatis*). The application of cross-adsorption and western blotting of rickettsial antigens revealed that *R. rickettsii* is not the only cause of rickettsiosis in the study area; *R. parkeri* and *R. amblyommatis* are also common agents causing spotted fever (Table 1).

Travelers returning from different parts of the world are exposed to diseases caused by pathogenic *Rickettsia* endemic to the country. In countries such as the United States, the most frequent rickettsiosis among travelers (90% of cases) is African tick-bite fever, caused by *Rickettsia africae*. However, in the USA, there are no specific tests for *R. africae*, and most available tests use antigens from *R. conorii* or *R. rickettsii* [63]. Although the antigens are different, cross-reactivity between the rickettsiae of the spotted fever group is evident, making them helpful in diagnosing *R. rickettsii* infection [34].

Regarding serological cross-reactivity in animals, we did not find antibodies that cross-reacted with other species of SFG Rickettsia at 16 DPI. These data contrast with those of Simser et al. [52], who reported the cross-reactivity of *R. monacensis*, *Rickettsia peacockii*, *R. rickettsii*, and *Rickettsia helvetica* in Syrian hamsters. In that study, sera were collected 50 days post-infection, and antibody titers between 1:64 and 1:512 were obtained. In our study, anti-*Ca*. R. colombiensis IgG antibody titers ranged from 1:64 to 1:1028, and there was no cross-reactivity between *R. rickettsii* and *R. parkeri* species. A possible explanation for these results is that the infection time in our study was 16 days post-infection (DPI). The appearance of heterologous antibodies that react with other Rickettsia species may be possible, thereby prolonging the post-infection period.

A study on the mechanisms of cell-mediated immunity demonstrated that guinea pigs were protected against *R. rickettsii* infection by *R. rhipicephali* [64]. The study reported that 100% of the animals in the three groups infected with *R. rhipicephali* at different doses (low: 7.5 × 10^2^ PFU, medium: 7.5 × 10^4^ PFU, and high: 7.5 × 10^6^ PFU) had antibodies that cross-reacted with *R. rickettsii* from 10 DPI. These results contrast with those reported in our study, which no cross-reactivity was observed with other SFG species (*R. rickettsii* and *R. parkeri*). We believe that several variables, such as the animal model used (Syrian hamster) and the time of exposure to infection (16 DPI), partly explain the difference in the results.

The reported experience with other *Rickettsia* species that were initially described in the vector and did not cause disease in humans but were later established to be pathogenic demonstrates that further studies are needed, such as experimental transmission trials by ticks to mammalian hosts and antibody titer assessments in subjects, which will be essential to determine the pathogenic potential of *Ca*. R. colombiensis. One of the weaknesses of our study was that we did not evaluate the sera with serum cross-adsorption or western blotting to establish the species of *Rickettsia* involved in the infection of the sera used. Another possible weakness of this study is the short duration (16 days) of infection with *Ca*. R. colombiensis in hamsters. Increasing the reaction time increased the number of antibodies, including heterologous antibodies.

In conclusion, *Ca*. R. colombiensis caused a subclinical infection in Syrian hamsters, suggesting the possibility of infecting other mammalian hosts. However, the clinical, pathological, and molecular findings are inconclusive regarding the establishment of *Ca*. R. colombiensis is a pathogenic species of SFG. As previously known, rickettsial cross-reactivity makes it difficult to diagnose a specific rickettsiosis, although it could be useful for confirming rickettsial syndrome.

## Supporting information

**S1 Fig.** S1 fig. Histopathological analysis of different hamsters tissues. A and B correspondent to lesions of a genetic background found normally in kidney and liver. A) lesions in kidney: focal membranoproliferative glomerulopathy, moderate multifocal lymphocytosis in the interstitial tubule. B) mild multifocal periportal plasmacytosis in the liver. C to H correspondent to histopathological analysis animal control tissues.

**S1 Fig A MPI 16 ID Kidney**

**S1 Fig B MPI 16 ID Liver**

**S1 Fig C CONTROL16-IP Liver**

**S1 Fig D CONTROL16-IP Lung**

**S1 Fig E CONTROL16-IP Brain**

**S1 Fig F CONTROL16-IP Spleen**

**S1 Fig G CONTROL16-IP Aorta**

**S1 Fig H CONTROL16-IP Kidney**

## Author contributions

**Conceptualization:** Jorge Miranda, Alejandra García, Salim Mattar

**Data curation:** Jorge Miranda, Alejandra García.

**Formal analysis:** Jorge Miranda, Alejandra García.

**Investigation:** Jorge Miranda, Alejandra García, Sonia Santibáñez, Cristina Cervera-Acevedo.

**Methodology:** Jorge Miranda, Alejandra García, Salim Mattar.

**Project administration:** Jorge Miranda, Salim Mattar.

**Resources:** Salim Mattar, Sonia Santibáñez, Aránzazu Portillo, José Oteo.

**Software:** Jorge Miranda.

**Supervision:** Jorge Miranda, Salim Mattar.

**Validation:** Jorge Miranda, Alejandra García, Sonia Santibáñez, Cristina Cervera-Acevedo, Salim Mattar.

**Visualization:** Jorge Miranda, Alejandra García, Salim Mattar.

**Writing – original draft:** Jorge Miranda, Salim Mattar.

**Writing – review & editing:** Jorge Miranda, Salim Mattar, Alejandra García, Sonia Santibáñez, Cristina Cervera-Acevedo, Aránzazu Portillo, José Oteo.

## Data Availability

the authors declare that all data underlying their findings fully available without restriction.

## Acknowledgments

Jorge Miranda acknowledge to Doctorado en Microbiología y Salud Tropical program and the Universidad de Córdoba. Also thanks to program Becas de Excelencia Doctoral del Bicentenario of Ministerio de Ciencias Tecnología e Innovación (BPIN 2019000100032) and Sistema General de Regalías de Córdoba for their support.

## References

1. Parola P, Paddock CD, Socolovschi C, Labruna MB, Mediannikov O, Kernif T, et al. Update on tick-borne rickettsioses around the world: a geographic approach. Clin Microbiol Rev. 2013;26: 657–702. doi: 10.1128/CMR.00032-13.

2. Sahni A, Fang R, Sahni SK, Walker DH. Pathogenesis of rickettsial diseases: pathogenic and immune mechanisms of an endotheliotropic infection. Annual Review of Pathology: Mechanisms of Disease. 2019;14: 127–152.

3. Sit B, Lamason RL. Pathogenic Rickettsia spp. as emerging models for bacterial biology. J Bacteriol. 2024;206: e0040423–23. Epub 2024 Feb 5. doi: 10.1128/jb.00404-23.

4. Helminiak L, Mishra S, Kim HK. Pathogenicity and virulence of Rickettsia. Virulence. 2022;13: 1752–1771.

5. Weinert LA, Werren JH, Aebi A, Stone GN, Jiggins FM. Evolution and diversity of Rickettsia bacteria. BMC biology. 2009;7: 1–15.

6. Salje J. Cells within cells: Rickettsiales and the obligate intracellular bacterial lifestyle. Nature Reviews Microbiology. 2021;19: 375–390.

7. Parola P, Musso D, Raoult D. Rickettsia felis: the next mosquito-borne outbreak? The Lancet Infectious Diseases. 2016;16: 1112–1113.

8. Gillespie JJ, Williams K, Shukla M, Snyder EE, Nordberg EK, Ceraul SM, et al. Rickettsia phylogenomics: unwinding the intricacies of obligate intracellular life. PloS one. 2008;3: e2018.

9. Snellgrove AN, Krapiunaya I, Scott P, Levin ML. Assessment of the pathogenicity of Rickettsia amblyommatis, Rickettsia bellii, and Rickettsia montanensis in a guinea pig model. Vector-Borne and Zoonotic Diseases. 2021;21: 232–241.

10. Tomassone L, Portillo A, Nováková M, De Sousa R, Oteo JA. Neglected aspects of tick-borne rickettsioses. Parasites & vectors. 2018;11: 1–11.

11. Paddock CD, Sumner JW, Comer JA, Zaki SR, Goldsmith CS, Goddard J, et al. Rickettsia parkeri: a newly recognized cause of spotted fever rickettsiosis in the United States. Clinical Infectious Diseases. 2004;38: 805–811.

12. Raoult D, Berbis P, Roux V, Xu W, Maurin M. A new tick-transmitted disease due to Rickettsia slovaca. Lancet. 1997;350: 112–113. doi: 10.1016/S0140-6736(05)61814-4.

13. Portillo A, Ibarra V, Santibáñez S, Pérez-Martínez L, Blanco JR, Oteo JA. Genetic characterisation of ompA, ompB and gltA genes from Candidatus Rickettsia rioja. Clinical Microbiology and Infection. 2009;15: 307–308.

14. Probert WS, Haw MP, Nichol AC, Glaser CA, Park SY, Campbell LE, et al. Newly Recognized Spotted Fever Group Rickettsia as Cause of Severe Rocky Mountain Spotted Fever– Like Illness, Northern California, USA. Emerging Infectious Diseases. 2024;30: 1344.

15. Salje J, Weitzel T, Newton PN, Varghese GM, Day N. Rickettsial infections: A blind spot in our view of neglected tropical diseases. PLoS Neglected Tropical Diseases. 2021;15: e0009353.

16. Zhang Y, Sun Y, Chen J, Teng A, Wang T, Li H, et al. Mapping the global distribution of spotted fever group rickettsiae: a systematic review with modelling analysis. The Lancet Digital Health. 2023;5: e5–e15.

17. Miranda J, Portillo A, Oteo JA, Mattar S. Rickettsia sp. strain colombianensi (Rickettsiales: Rickettsiaceae): a new proposed Rickettsia detected in Amblyomma dissimile (Acari: Ixodidae) from iguanas and free-living larvae ticks from vegetation. J Med Entomol. 2012;49: 960-965.

18. Imaoka K, Kaneko S, Tabara K, Kusatake K, Morita E. The First Human Case of Rickettsia tamurae Infection in Japan. Case Rep Dermatol. 2011;3: 68–73. doi: 10.1159/000326941.

19. Jado I, Oteo JA, Aldamiz M, Gil H, Escudero R, Ibarra V, et al. Rickettsia monacensis and human disease, Spain. Emerg Infect Dis. 2007;13: 1405-1407. doi: 10.3201/eid1309.060186.

20. Oren A, Garrity GM, Parker CT, Chuvochina M, Trujillo ME. Lists of names of prokaryotic Candidatus taxa. Int J Syst Evol Microbiol. 2020;70: 3956–4042. doi: 10.1099/ijsem.0.003789.

21. Cotes-Perdomo A, Cárdenas-Carreño J, Hoyos J, González C, Castro LR. Molecular detection of Candidatus Rickettsia colombianensi in ticks (Acari, Ixodidae) collected from herpetofauna in San Juan de Carare, Colombia. International Journal for Parasitology: Parasites and Wildlife. 2022;19: 110–114.

22. Lerma LS, Cogollo VC, Velilla SM, Gonzalez IR, Atehortua ADM, Peñuela DFC. First detection of Candidatus Rickettsia colombianensi in the State of Meta, Colombia. Revista Habanera de Ciencias Médicas. 2019;18: 487–499.

23. Martínez-Sánchez ET, Cardona-Romero M, Ortiz-Giraldo M, Tobón-Escobar WD, Moreno-López D, Ossa-López PA, et al. Rickettsia spp. in ticks (Acari: Ixodidae) from wild birds in Caldas, Colombia. Acta Trop. 2021;213: 105733.

24. Rivera-Páez FA, Martins TF, Ossa-López PA, Sampieri BR, Camargo-Mathias MI. Detection of Rickettsia spp. in ticks (Acari: Ixodidae) of domestic animals in Colombia. Ticks and tick-borne diseases. 2018;9: 819-823.

25. Santodomingo A, Cotes-Perdomo A, Foley J, Castro LR. Rickettsial infection in ticks (Acari: Ixodidae) from reptiles in the Colombian Caribbean. Ticks and tick-borne diseases. 2018;9: 623-628.

26. Bermúdez S, Martínez-Mandiche J, Domínguez L, Gonzalez C, Chavarria O, Moreno A, et al. Diversity of Rickettsia in ticks collected from wild animals in Panama. Ticks and Tick-borne Diseases. 2021;12: 101723.

27. Luz HR, Silva-Santos E, Costa-Campos CE, Acosta I, Martins TF, Muñoz-Leal S, et al. Detection of Rickettsia spp. in ticks parasitizing toads (Rhinella marina) in the northern Brazilian Amazon. Experimental and Applied Acarology. 2018;75: 309–318.

28. Ogrzewalska M, Machado C, Rozental T, Forneas D, Cunha LE, De Lemos E. Microorganisms in the ticks Amblyomma dissimile Koch 1844 and Amblyomma rotundatum Koch 1844 collected from snakes in Brazil. Med Vet Entomol. 2019;33: 154–161.

29. Romero L, Costa FB, Labruna MB. Ticks and tick-borne Rickettsia in El Salvador. Experimental and Applied Acarology. 2021;83: 545–554.

30. Sánchez-Montes S, Isaak-Delgado AB, Guzmán-Cornejo C, Rendón-Franco E, Muñoz-García CI, Bermúdez S, et al. Rickettsia species in ticks that parasitize amphibians and reptiles: Novel report from Mexico and review of the worldwide record. Ticks and tick-borne diseases. 2019;10: 987–994.

31. Cotes-Perdomo A, Santodomingo A, Castro LR. Hemogregarine and Rickettsial infection in ticks of toads from northeastern Colombia. Int J Parasitol Parasites Wildl. 2018;7: 237–242. doi: 10.1016/j.ijppaw.2018.06.003.

32. Miranda J, Violet-Lozano L, Barrera S, Mattar S, Monsalve-Buriticá S, Rodas J, et al. Candidatus Rickettsia colombianensi in ticks from reptiles in Córdoba, Colombia. Veterinary world. 2020;13: 1764.

33. Perlman SJ, Hunter MS, Zchori-Fein E. The emerging diversity of Rickettsia. Proc Biol Sci. 2006;273: 2097–2106. doi: 10.1098/rspb.2006.3541.

34. Oteo JA, Nava S, Sousa Rd, Mattar S, Venzal JM, Abarca K, et al. Guías Latinoamericanas de la RIICER para el diagnóstico de las rickettsiosis transmitidas por garrapatas. Revista chilena de infectología. 2014;31: 54–65.

35. Gimenez DF. Staining Rickettsiae in Yolk-Sac Cultures. Stain Technol. 1964;39: 135–140. doi: 10.3109/10520296409061219.

36. Labruna MB, Whitworth T, Horta MC, Bouyer DH, McBride JW, Pinter A, et al. Rickettsia species infecting Amblyomma cooperi ticks from an area in the state of São Paulo, Brazil, where Brazilian spotted fever is endemic. J Clin Microbiol. 2004;42: 90–98.

37. Kelly PJ, Raoult D, Mason PR. Isolation of spotted fever group rickettsias from triturated ticks using a modification of the centrifugation-shell vial technique. Trans R Soc Trop Med Hyg. 1991;85: 397–398. doi: 10.1016/0035-9203(91)90303-g.

38. Feng WC, Waner JL. Serological cross-reaction and cross-protection in guinea pigs infected with Rickettsia rickettsii and Rickettsia montana. Infect Immun. 1980;28: 627–629. doi: 10.1128/iai.28.2.627-629.1980.

39. Brustolin JM, da Silva Krawczak F, Alves MEM, Weiller MA, de Souza CL, Rosa FB, et al. Experimental infection in Cavia porcellus by infected Amblyomma ovale nymphs with Rickettsia sp. (Atlantic rainforest strain). Parasitol Res. 2018;117: 713–720. doi: 10.1007/s00436-017-5741-2.

40. Soares JF, Soares HS, Barbieri AM, Labruna MB. Experimental infection of the tick Amblyomma cajennense, Cayenne tick, with Rickettsia rickettsii, the agent of Rocky Mountain spotted fever. Med Vet Entomol. 2012;26: 139–151. doi: 10.1111/j.1365-2915.2011.00982.x.

41. Paris DH, Dumler JS. State of the art of diagnosis of rickettsial diseases: the use of blood specimens for diagnosis of scrub typhus, spotted fever group rickettsiosis, and murine typhus. Curr Opin Infect Dis. 2016;29: 433–439.

42. Horta MC, Labruna MB, Sangioni LA, Vianna MC, Gennari SM, Galvão MA, et al. Prevalence of antibodies to spotted fever group rickettsiae in humans and domestic animals in a Brazilian spotted fever-endemic area in the state of São Paulo, Brazil: serologic evidence for infection by Rickettsia rickettsii and another spotted fever group Rickettsia. Am J Trop Med Hyg. 2004;71: 93-97.

43. Portillo A, De Sousa R, Santibáñez S, Duarte A, Edouard S, Fonseca IP, et al. Guidelines for the Detection of Rickettsia spp. Vector-Borne and Zoonotic Diseases. 2017;17: 23–32.

44. Stewart AG, Stewart AG. An update on the laboratory diagnosis of Rickettsia spp. infection. Pathogens. 2021;10: 1319.

45. Roux V, Fournier PE, Raoult D. Differentiation of spotted fever group rickettsiae by sequencing and analysis of restriction fragment length polymorphism of PCR-amplified DNA of the gene encoding the protein rOmpA. J Clin Microbiol. 1996;34: 2058–2065. doi: 10.1128/jcm.34.9.2058-2065.1996.

46. Kumar S, Stecher G, Li M, Knyaz C, Tamura K. MEGA X: Molecular Evolutionary Genetics Analysis across Computing Platforms. Mol Biol Evol. 2018;35: 1547–1549. doi: 10.1093/molbev/msy096.

47. National Toxicology Program. Nonneoplastic Lesion Atlas [cited 13 December 2024]. Available: https://ntp.niehs.nih.gov/atlas/nnl.

48. Miao J, Chard LS, Wang Z, Wang Y. Syrian hamster as an animal model for the study on infectious diseases. Frontiers in immunology. 2019;10: 2329.

49. Stokes JV, Walker DH, Varela-Stokes AS. The guinea pig model for tick-borne spotted fever rickettsioses: A second look. Ticks and tick-borne diseases. 2020;11: 101538.

50. Feng H, Wen J, Walker DH. Rickettsia australis infection: a murine model of a highly invasive vasculopathic rickettsiosis. The American journal of pathology. 1993;142: 1471.

51. Londoño AF, Mendell NL, Walker DH, Bouyer DH. A biosafety level-2 dose-dependent lethal mouse model of spotted fever rickettsiosis: Rickettsia parkeri Atlantic Rainforest strain. PLoS neglected tropical diseases. 2019;13: e0007054.

52. Simser JA, Palmer AT, Fingerle V, Wilske B, Kurtti TJ, Munderloh UG. Rickettsia monacensis sp. nov., a spotted fever group Rickettsia, from ticks (Ixodes ricinus) collected in a European city park. Appl Environ Microbiol. 2002;68: 4559–4566.

53. McDade JE. Evidence supporting the hypothesis that rickettsial virulence factors determine the severity of spotted fever and typhus group infections. Ann N Y Acad Sci. 1990;590: 20–26. doi: 10.1111/j.1749-6632.1990.tb42202.x.

54. Blanton LS, Mendell NL, Walker DH, Bouyer DH. “Rickettsia amblyommii” induces cross protection against lethal Rocky Mountain spotted fever in a guinea pig model. Vector-Borne and Zoonotic Diseases. 2014;14: 557–562.

55. Burgdorfer W, Hayes SF, Thomas LA, Lancaster JL. New spotted fever group Rickettsia from the lone star tick, Amblyomma americanum. Rickettsiae and rickettsial diseases / edited by W. Burgdorfer; RL Anacker. 1981.

56. Rivas JJ, Moreira-Soto A, Alvarado G, Taylor L, Calderón-Arguedas O, Hun L, et al. Pathogenic potential of a Costa Rican strain of ‘Candidatus Rickettsia amblyommii’in guinea pigs (Cavia porcellus) and protective immunity against Rickettsia rickettsii. Ticks and Tick-borne Diseases. 2015;6: 805–811.

57. Philip* RN, Casper EA, Anacker RL, Cory J, Hayes SF, Burgdorfer W, et al. Rickettsia bellii sp. nov.: a tick-borne rickettsia, widely distributed in the United States, that is distinct from the spotted fever and typhus biogroups. Int J Syst Evol Microbiol. 1983;33: 94–106.

58. Horta MC, Sabatini GS, Moraes-Filho J, Ogrzewalska M, Canal RB, Pacheco RC, et al. Experimental infection of the opossum Didelphis aurita by Rickettsia felis, Rickettsia bellii, and Rickettsia parkeri and evaluation of the transmission of the infection to ticks Amblyomma cajennense and Amblyomma dubitatum. Vector-Borne and Zoonotic Diseases. 2010;10: 959–967.

59. de Sousa R, Dos Santos ML, Cruz C, Almeida V, Garrote AR, Ramirez F, et al. Rare case of rickettsiosis caused by Rickettsia monacensis, Portugal, 2021. Emerging Infectious Diseases. 2022;28: 1068.

60. Delisle J, Mendell NL, Stull-Lane A, Bloch KC, Bouyer DH, Moncayo AC. Human infections by multiple spotted fever group rickettsiae in Tennessee. Am J Trop Med Hyg. 2016;94: 1212.

61. Quintero Velez JC, Aguirre-Acevedo DC, Rodas JD, Arboleda M, Troyo A, Vega Aguilar F, et al. Epidemiological characterization of incident cases of Rickettsia infection in rural areas of Uraba region, Colombia. PLoS Negl Trop Dis. 2018;12: e0006911. doi: 10.1371/journal.pntd.0006911.

62. Vaughn MF, Delisle J, Johnson J, Daves G, Williams C, Reber J, et al. Seroepidemiologic study of human infections with spotted fever group rickettsiae in North Carolina. J Clin Microbiol. 2014;52: 3960–3966.

63. Biggs HM. Diagnosis and management of tickborne rickettsial diseases: Rocky Mountain spotted fever and other spotted fever group rickettsioses, ehrlichioses, and anaplasmosis—United States. MMWR.Recommendations and Reports. 2016;65.

64. Gage KL, Jerrells TR. Demonstration and partial characterization of antigens of Rickettsia rhipicephali that induce cross-reactive cellular and humoral immune responses to Rickettsia rickettsii. Infect Immun. 1992;60: 5099–5106.

